# Public interest in postural orthostatic tachycardia syndrome in the United Kingdom, 2004–2026: a Google Trends infodemiology study

**DOI:** 10.64898/2026.08.21.26361021

**Authors:** Richard G Bogle, Cecilia ME Bogle

## Abstract

**Background:** Public and clinical attention to postural orthostatic tachycardia syndrome (POTS) has increased, particularly since the COVID-19 pandemic. We quantified changes in United Kingdom Google search interest and examined whether searches increasingly used diagnostic and self-assessment language.

**Methods:** We extracted monthly Google Trends relative search volume (RSV; 0–100) for the Health-category search term “Pots syndrome” in the United Kingdom from January 2004 through July 2026. Five extraction attempts were made; two returned complete, identical monthly series and were retained. Prespecified eras were summarised and an exploratory interrupted time-series model at March 2020 used ordinary least squares with Newey–West heteroskedasticity- and autocorrelation-consistent standard errors (12 lags). Comparator searches included conventional orthostatic diagnoses, POTS diagnostic terms, associated conditions and YouTube searches.

**Results:** The primary series comprised 271 complete months. Mean RSV increased from 18.6 during 2015–2019 to 64.8 during 2022–2023 (3.49-fold) and remained 50.6 during January 2024–July 2026 (2.73-fold above baseline). Search interest peaked in October 2022 (RSV 100); July 2026 RSV was 57. The interrupted time-series model estimated an immediate March 2020 level increase of 21.8 points (95% CI 2.8–40.7; p=0.024), while the slope change was not statistically supported (0.069 points/month, 95% CI −0.299 to 0.438; p=0.713). Searches for “POTS symptoms”, “POTS test” and “POTS heart rate” increased more steeply than the general term, although low baseline volumes made fold changes unstable.

**Conclusions:** UK Google search interest in POTS rose before 2020, increased sharply after the pandemic began, and remained substantially above its prepandemic baseline. The results demonstrate a sustained change in public attention, not disease incidence or social-media causation. The growth of symptom- and testing-oriented searches is compatible with increased diagnostic self-investigation and warrants linkage to referral, diagnosis and social-media exposure data.

## Introduction

Postural orthostatic tachycardia syndrome (POTS) is a chronic syndrome characterised by orthostatic intolerance with an excessive sustained increase in heart rate on standing, in the absence of orthostatic hypotension or another condition sufficient to explain the tachycardia. Contemporary criteria require a sustained heart-rate increase of at least 30 beats/min in adults, or 40 beats/min at ages 12–19 years, within 10 minutes of upright posture, together with chronic posture-dependent symptoms and exclusion of alternative causes.^1–3^ The haemodynamic threshold is nevertheless protocol-sensitive and is not, by itself, a disease-specific biomarker.□

Recognition of POTS has historically been poor. A large community survey reported a median diagnostic delay of 24 months and frequent previous misdiagnosis.□ Increased awareness may therefore correct genuine unmet need. At the same time, POTS comprises heterogeneous symptom and mechanistic profiles, many symptoms are nonspecific, and orthostatic tachycardia can occur secondary to acute illness, hypovolaemia, anaemia, medication, endocrine disease or deconditioning.^1–3^ These features create potential both for underdiagnosis and for premature attribution of diverse symptoms to a single label.

The COVID-19 pandemic introduced an additional biological and informational discontinuity. Autonomic symptoms and POTS phenotypes have been described after SARS-CoV-2 infection, but estimates are affected by referral selection, varying diagnostic definitions and heterogeneous testing.□–□ Concurrently, online patient communities, consumer heart-rate wearables and searchable diagnostic thresholds have made POTS unusually visible and personally measurable. Qualitative research indicates that diagnostic labels can shape illness identity, symptom interpretation and management behaviours among people seeking a POTS diagnosis.□ This is consistent with the sociological concept of “looping”, in which classifications alter how people understand and present themselves, thereby changing the population to which the classification is subsequently applied.^1^□

Google Trends provides sampled, anonymised and normalised measures of population search interest. It cannot establish prevalence, motivation or causal exposure, but it can identify when public attention changes and which information is sought.^11–13^ We therefore examined UK Google search interest in POTS from 2004 to 2026. We hypothesised that interest would show a sustained increase after March 2020 and that symptom-, testing- and heart-rate-oriented queries would grow disproportionately. The study was designed to distinguish evidence of changing attention from stronger, unsupported claims about disease incidence or social-media causation.

## Methods

### Study design and data source

We conducted an infodemiological time-series study using Google Trends. Google Trends reports relative search volume (RSV), normalised to the proportion of all Google searches in the selected geography and period and rescaled from 0 to 100. A value of 100 denotes the point of greatest relative interest within a request; zero can indicate insufficient sampled volume and does not prove that no searches occurred.^11^

Data were collected on 21 August 2026. The primary request used the literal search term “Pots syndrome”, geography United Kingdom, category Health, search surface Web Search and period January 2004 to the latest available month. August 2026 was incomplete and was excluded, leaving January 2004–July 2026. The Health category was chosen to reduce contamination from searches concerning cooking or plant pots.

### Reproducibility and repeated extraction

Five identical primary extraction attempts were made because Google Trends uses sampled data and repeated results may vary. Two attempts returned complete monthly tables; three rendered without a complete data table and were retained in the audit log but were not treated as zero. The two complete series were identical. The prespecified processed value was the monthly median across valid repetitions, with minimum and maximum retained. All request URLs, timestamps, rendered-row counts and failures were recorded in a supplementary workbook.

### Comparison searches

Prespecified comparison batches retained “Pots syndrome” as a common anchor. Conventional diagnostic comparators were “orthostatic hypotension”, “vasovagal syncope”, “inappropriate sinus tachycardia” and “supraventricular tachycardia”. Associated-condition searches were “Ehlers-Danlos syndrome”, “mast cell activation syndrome”, “ME/CFS” and “long COVID”. Diagnostic-language searches were “POTS symptoms”, “POTS diagnosis”, “POTS test” and “POTS heart rate”. Testing and monitoring searches included “tilt table test”, “standing test”, “Apple Watch heart rate” and “heart rate monitor”. A YouTube Search sensitivity analysis included POTS and dysautonomia terms. Each comparison series was interpreted only within its own Google Trends request because scaling changes across requests.

Top and rising related-query panels were sought for six historical windows: 2004–2009, 2010–2014, 2015–February 2020, March 2020–2021, 2022–2023 and 2024–July 2026. Google reported insufficient data in the relevant panels; these data were therefore not analysed.

### Statistical analysis

We calculated the mean, median, range and number of months for six prespecified eras. The primary comparative baseline was January 2015–December 2019. Ratios of mean RSV in 2022–2023 and January 2024–July 2026 to this baseline were calculated descriptively.

An exploratory segmented ordinary least-squares regression used March 2020 as a prespecified interruption. The model included continuous month, an indicator for the post-March-2020 period and months elapsed after the interruption. Newey–West heteroskedasticity- and autocorrelation-consistent standard errors used 12 monthly lags. Coefficients represent the prepandemic monthly slope, immediate level change and change in slope. Because Google Trends values are bounded, normalised and sampled, this model was treated as descriptive rather than causal. Analyses were performed in Python 3. No adjustment was made for multiple exploratory comparisons.

### Ethics and reporting

Only publicly available, aggregated and anonymised search-interest data were used. No individual-level data, personal identifiers or user histories were accessed; research ethics approval and participant consent were therefore not sought. Reporting was informed by established recommendations for reproducible Google Trends research.^12,13^

## Results

### Primary time series

The primary dataset comprised 271 complete monthly observations from January 2004 through July 2026. Search interest was absent or below Google’s reporting threshold through most of 2004–2008; the first non-zero monthly value occurred in April 2009. Mean RSV was 7.8 during 2010–2014 and 18.6 during January 2015–February 2020. It rose to 37.5 during March 2020–December 2021 and 64.8 during 2022–2023. The peak occurred in October 2022 (RSV 100). Mean RSV subsequently declined but remained 50.6 during January 2024–July 2026; the final monthly value was 57.

Relative to the January 2015–December 2019 mean of 18.6, mean RSV was 3.49-fold higher in 2022– 2023 and 2.73-fold higher in January 2024–July 2026. Thus, the post-peak series did not return to its prepandemic baseline (Table 1; Figure 1).

**Table 1.** Relative Google search interest for “Pots syndrome” by prespecified era.

| Period | Months | Mean RSV | Median RSV | Minimum | Maximum |
| --- | --- | --- | --- | --- | --- |
| 2004-2009 | 72 | 0.1 | 0.0 | 0 | 6 |
| 2010-2014 | 60 | 7.8 | 7.0 | 0 | 21 |
| 2015-Feb2020 | 62 | 18.6 | 18.0 | 11 | 34 |
| Mar2020-2021 | 22 | 37.5 | 40.5 | 15 | 54 |
| 2022-2023 | 24 | 64.8 | 60.0 | 40 | 100 |
| 2024-Jul2026 | 31 | 50.6 | 50.0 | 41 | 72 |

RSV, relative search volume. The final period ends July 2026; August 2026 was incomplete and excluded.

**Figure 1.**
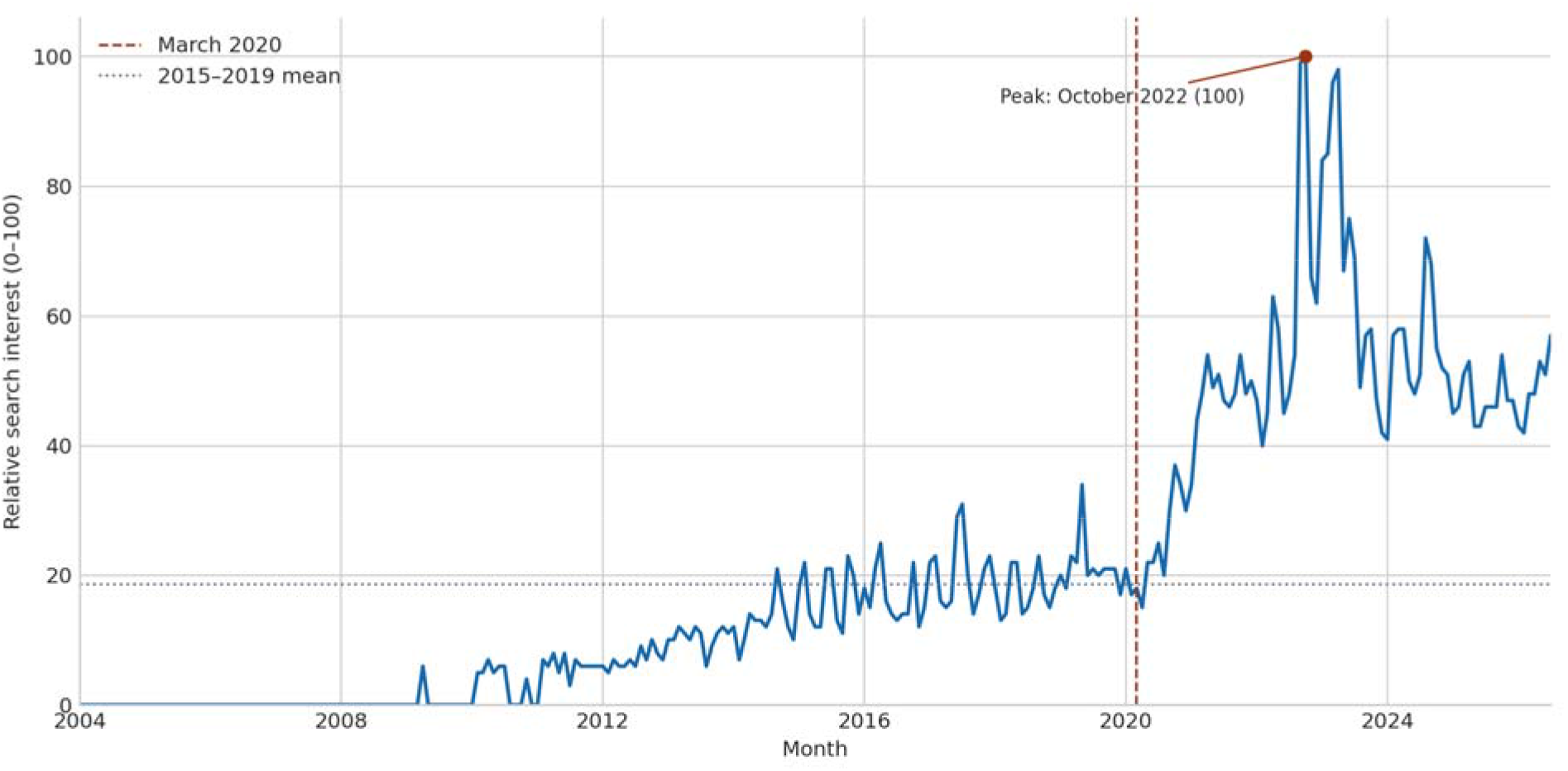
Monthly UK Google search interest in “Pots syndrome”, January 2004–July 2026. *The dashed line marks March 2020; the dotted line is the January 2015–December 2019 mean. Google Trends values are normalised within the request and do not represent absolute search counts*.

### Interrupted time-series analysis

The estimated prepandemic increase was 0.137 RSV points per month (95% CI 0.118–0.156; p<0.001). At March 2020, the model estimated an immediate level increase of 21.8 points (95% CI 2.8–40.7; p=0.024). The subsequent slope change was 0.069 points per month (95% CI -0.299 to 0.438; p=0.713). The absence of a statistically supported slope change indicates that the dominant modelled discontinuity was a level shift superimposed on an established upward trend, rather than evidence that the pandemic initiated interest from zero.

### Comparator and diagnostic-language searches

Among conventional diagnostic comparators, the 2022–2023 to 2015–2019 mean ratio was 1.97 for orthostatic hypotension, 1.86 for vasovagal syncope and 1.24 for supraventricular tachycardia, compared with 3.49 for POTS. Inappropriate sinus tachycardia increased 5.15-fold but had a very low baseline RSV (0.55), making its ratio unstable. By 2024–July 2026, POTS remained 2.73-fold above baseline, while supraventricular tachycardia was 1.26-fold above baseline (Table 2).

**Table 2.** Mean relative search interest in selected comparison terms.

| Search term | 2015–2019 | 2022–2023 | 2024–Jul 2026 | 2024–Jul 2026 / baseline |
| --- | --- | --- | --- | --- |
| Pots syndrome | 18.57 | 64.75 | 50.65 | 2.73 |
| orthostatic hypotension | 10.08 | 19.88 | 27.19 | 2.70 |
| vasovagal syncope | 18.47 | 34.38 | 42.61 | 2.31 |
| inappropriate sinus tachycardia | 0.55 | 2.83 | 3.23 | 5.87 |
| supraventricular tachycardia | 11.45 | 14.17 | 14.45 | 1.26 |
| POTS symptoms | 5.12 | 27.62 | 76.32 | 14.92 |
| POTS test | 0.52 | 4.38 | 8.48 | 16.42 |
| POTS heart rate | 0.12 | 2.67 | 7.00 | 60.00 |

*Values are scaled within their comparison request. Fold changes for low-volume terms are unstable and should not be compared as estimates of population incidence.*

Within the diagnostic-language request, mean RSV for “POTS symptoms” increased from 5.1 in 2015– 2019 to 27.6 in 2022–2023 and 76.3 in 2024–July 2026. “POTS test” increased from 0.52 to 4.38 and 8.48, respectively; “POTS heart rate” increased from 0.12 to 2.67 and 7.00. These patterns suggest increasing interest in symptom recognition and self-assessment, but their large ratios partly reflect sparse baseline volumes and changing within-request scaling.

## Discussion

### Principal findings

UK Google search interest in POTS increased gradually for at least a decade before COVID-19, rose sharply after March 2020, peaked in late 2022 and remained substantially above its prepandemic baseline through July 2026. The sustained elevation after the peak is more informative than the isolated maximum: it suggests a durable change in public awareness or information-seeking rather than a single transient media event. The rise was larger than that for vasovagal syncope and supraventricular tachycardia, and diagnostic-language searches grew particularly rapidly.

These data do not establish why interest increased. At least four explanations remain plausible and may coexist: a true increase in post-infectious autonomic illness; improved professional and public recognition of previously missed POTS; expanded online patient advocacy and social-media exposure; and the increasing availability of consumer heart-rate data. The immediate level change at March 2020 is temporally compatible with both a biological effect of SARS-CoV-2 and a major change in health-information behaviour. Google Trends cannot separate these mechanisms.

### Diagnostic looping as a testable interpretation

POTS may be particularly susceptible to diagnostic looping because the category supplies both a broad symptom narrative and a numerical threshold that can be repeatedly examined with consumer wearables. Exposure to the category may help a person recognise genuine posture-dependent illness and obtain appropriate care. It may also increase attention to transient heart-rate changes, encourage unstandardised home testing and promote attribution of nonspecific fatigue, dizziness or cognitive symptoms to POTS before exclusions have been assessed. The resulting changes in monitoring, activity and clinical presentation could then influence the diagnosed population and subsequent descriptions of the syndrome.

The disproportionate growth of “POTS symptoms”, “POTS test” and “POTS heart rate” searches is compatible with this mechanism but does not demonstrate it. Search queries do not reveal whether users were patients, relatives, clinicians, journalists or researchers; whether social media prompted the search; whether a diagnosis followed; or whether that diagnosis was correct. “Looping” should therefore be treated as a hypothesis for individual-level prospective research, not as an explanation imposed on the present ecological data.

### Clinical and research implications

The findings support two simultaneous clinical priorities. First, increased awareness may reduce documented diagnostic delay and the inappropriate dismissal of genuine orthostatic illness.□ Second, greater attention increases the need to protect diagnostic specificity. Crossing a 30-beats/min threshold during an unstandardised stand or tilt test is not sufficient: symptoms must be chronic and posture-dependent, tachycardia sustained, orthostatic hypotension considered, and competing causes excluded.^1^–□

The next research step should link prospectively measured POTS-specific online exposure with pre-assessment diagnostic certainty, wearable checking behaviour, standardised haemodynamics and blinded clinical adjudication among consecutive referrals. This would distinguish improved case recognition from false-positive self-identification. Regional linkage of search interest with referral volumes, tilt testing and confirmed diagnoses could provide a complementary population-level analysis, although ecological confounding would remain.

### Strengths and limitations

Strengths include a 22-year national series, prespecified eras and comparison terms, explicit separation of search interest from incidence, repeated extraction attempts, retention of failed loads in an audit trail, exclusion of the incomplete current month and autocorrelation-robust uncertainty estimates. The full processed dataset and extraction metadata are supplied.

The study has substantial limitations. Google Trends provides sampled, normalised indices rather than absolute counts, demographic characteristics or individual trajectories. Normalisation means values from separate requests cannot be compared directly, and low-volume terms generate zeros or unstable ratios. Only two of five primary attempts returned complete tables, although the retained series were identical. The literal term “Pots syndrome” may miss searches using abbreviations or alternative wording, while the Health category may exclude relevant searches. Conversely, some ambiguity may remain. Related-query data were insufficient for the prespecified historical analysis. Search behaviour is influenced by internet access, platform use, news coverage and Google’s undisclosed sampling and categorisation processes.^11^– ^1^□

March 2020 was a prespecified interruption but not an isolated exposure: infection waves, long-COVID recognition, media coverage and changing healthcare access evolved over several years. The segmented linear model simplifies a nonlinear bounded series and should not be interpreted causally. Most importantly, Google searches cannot establish social-media exposure, symptom amplification, diagnostic accuracy, POTS prevalence or the biological incidence of post-COVID autonomic disease.

## Conclusions

Public search interest in POTS in the United Kingdom rose progressively before 2020, increased sharply after the start of the COVID-19 pandemic and remained approximately 2.7-fold above its 2015–2019 baseline through July 2026. Increasing searches for symptoms, tests and heart-rate criteria indicate a shift towards diagnostic self-investigation. These findings establish a sustained change in attention, not its cause. They justify prospective research linking online exposure and wearable monitoring to standardised haemodynamic phenotypes, diagnostic adjudication and clinical outcomes.

## Data Availability

All data produced are available online at the Githhub respository shown in the Data Availability Link.

https://github.com/richardbogle/POTS_Google_Trends_infodemiology_study

## Declarations

### Ethics approval

Ethical approval was not required because the study used only publicly available, aggregated and anonymised Google Trends data. No individual-level data or identifiers were accessed, and no participants were recruited.

### Data availability

The processed monthly series, comparison series, extraction log, verified request URLs and query specification are supplied as Supplementary Data S1 (Google_Trends_POTS_UK_Data_Package.xlsx). Google Trends source data remain subject to Google’s terms and sampling procedures.

### Code availability

Analysis code used to reproduce the descriptive and segmented analyses is available at https://github.com/richardbogle/POTS_Google_Trends_infodemiology_study.

### Funding

No specific funding was received for this study.

### Competing interests

The authors declare no competing interests.

### Author contributions

Richard G Bogle: conceptualisation, study design, methodology, investigation, project administration, writing – original draft, writing – review and editing, and guarantor. Cecilia ME Bogle: data extraction, formal analysis, writing – original draft, and writing – review and editing. Both authors approved the final manuscript and accept responsibility for the work.

### Use of artificial intelligence

OpenAI Codex was used to assist with data-processing code and initial language drafting. The authors verified the analysis, references, interpretation and final manuscript and take full responsibility for the submitted work.

## Acknowledgements

None.

## Notes

### Competing Interest Statement

The authors have declared no competing interest.

